# Longitudinal information extraction from clinical notes in rare diseases: an efficient approach with small language models

**DOI:** 10.64898/2026.03.30.26349388

**Authors:** Xiaomeng Wang, Carole Faviez, Marc Vincent, Judith Jeyafreeda Andrew, Emma Le Priol, Sophie Saunier, Bertrand Knebelmann, Rui Zhang, Nicolas Garcelon, Anita Burgun, Xiaoyi Chen

## Abstract

**Objectives:** Rare diseases often require longitudinal monitoring to characterise progression, yet much clinical information remains locked in unstructured electronic health records (EHRs). Efficient recovery of such data is critical for accurate prognostic modelling and clinical trial preparation. We aimed to develop and evaluate a small language model (SLM)-based pipeline for extracting longitudinal information from French clinical notes of patients with rare kidney diseases.

**Methods:** As a use case, we focused on serum creatinine, a key biomarker of kidney function. We analyzed 81 clinical notes comprising 200 measurements (triplet of date, value and unit). Four open-source SLMs (Mistral-7B, Llama-3.2-3B, Qwen3-4B, Qwen3-8B) were systematically tested with different prompting strategies in French and English. Outputs were post-processed to standardize formats and resolve inconsistencies, and performance was assessed across model size, prompting, language, and robustness to text duplication.

**Results:** All SLMs extracted structured triplets, with F1-scores ranging from 0.519 to 0.928 (Qwen3-8B), outperforming the rule-based baseline. Larger models generally performed better, while prompting strategy and language had modest effects across models. SLMs also showed variable robustness to duplicated content common in real-world EHR notes.

**Discussion:** Lightweight, locally deployable language models can accurately extract longitudinal biomarkers from unstructured clinical notes. Our findings highlight their practicality for rare diseases where data scarcity often limits task-specific model training.

**Conclusion:** SLMs provide a privacy-preserving and resource-efficient solution for recovering longitudinal biomarker trajectories from unstructured notes, offering potential to advance real-world research and patient care in rare kidney diseases.

**What is already known?:**

- Longitudinal monitoring is essential in rare kidney diseases, yet key biomarker data are often locked in unstructured clinical notes.
- Large language models (LLMs) have shown strong performance in clinical text processing tasks but face major challenges related to privacy, computational cost, and implementation feasibility in healthcare settings.
- Small language models (SLMs) are emerging as lightweight, locally deployable alternatives whose potential for clinical applications is increasingly recognized.

**What does this paper add?:**

- This study provides the first real-world evaluation of SLMs for extracting longitudinal biomarker measurements in rare kidney disease cohorts.
- It introduces and validates an efficient extraction pipeline that combines document preselection, SLM prompting, and post-processing to accurately retrieve biomarker measurements from French clinical notes.
- The findings show that SLM-based extraction can help mitigate data scarcity in rare diseases, thereby improving prognosis modeling and supporting clinical research.

## Introduction

Rare diseases, defined as conditions affecting fewer than 1 in 2000 people, encompass over 7000 distinct disorders and affect an estimated 300 million individuals worldwide (1). Their cumulative burden is substantial: around 80% are genetic, 70% manifest in childhood, 95% lack approved treatments, and one in three of affected children die before the age of five years. Patients with rare diseases often experience complex and fragmented care, with key clinical information documented in free-text narratives rather than structured fields. Real-world electronic health records (EHRs) represent therefore a valuable yet underexplored resource for studying disease natural history (2).

Approaches to information extraction from free text range from rule-based systems to deep learning-based natural language processing (NLP) algorithms (3). Most NLP methods rely on task-specific training, which remains challenging in rare diseases due to the scarcity of annotated data. More recently, large language models (LLMs) have been studied across various clinical NLP tasks, including named entity recognition, information extraction, and summarization (4–8). Prompting enables the use of these pre-trained models even in data-scarce domains, reducing the need for extensive fine-tuning. However, deploying LLMs in healthcare settings poses challenges related to data privacy and computational cost (9).

Small language models (SLMs), also referred to as lightweight LLMs, offer a practical alternative for deployment in resource-constrained environments and for targeted information extraction in rare disease research (10,11). SLMs are typically defined as models with fewer than 10 billion parameters (12–14). Examples include Mistral-7B and smaller variants within the LLaMA and Qwen families, which originate from larger foundation models through pruning, quantization, or knowledge distillation (15–17). Despite their potential, to our knowledge, no studies have investigated whether SLMs can extract structured longitudinal biomarker data from real-world clinical notes, particularly in multilingual contexts such as French.

Rare kidney diseases account for a substantial proportion of adult and nearly all paediatric chronic kidney diseases (CKD) and renal failure. However, unlike common CKD caused by hypertension or diabetes, their progression patterns remain poorly characterized due to limited data and high heterogeneity in disease courses (18). Reconstructing longitudinal kidney function trajectories across these populations is therefore essential to improve prognosis, guide patient care, and design future therapeutic trials. Kidney function is typically monitored using estimated glomerular filtration rate (eGFR), derived from serum creatinine, which remains a key prognostic biomarker (19). Renal ciliopathies, a group of rare genetic disorders causing CKD and renal failure in adolescence or early adulthood, remain particularly under-studied because structured databases contain limited longitudinal data (20). Our previous work showed that clinical notes capture numerous creatinine measurements absent from these databases, thereby extending the temporal coverage of follow-up and improving the accuracy of longitudinal analyses (21).

However, the extraction process still required manual review, underscoring the need for scalable and automated solutions.

To address this gap, we developed and evaluated an SLM-based pipeline to extract longitudinal serum creatinine measurements from French clinical notes. Using real-world data from a national rare disease reference center, we benchmarked multiple SLMs under different prompting strategies and languages, demonstrating the feasibility of accurate and privacy-preserving extraction to support the longitudinal monitoring of kidney function.

## Methods

Figure 1 summarizes the overall study workflow.

**Figure 1.**
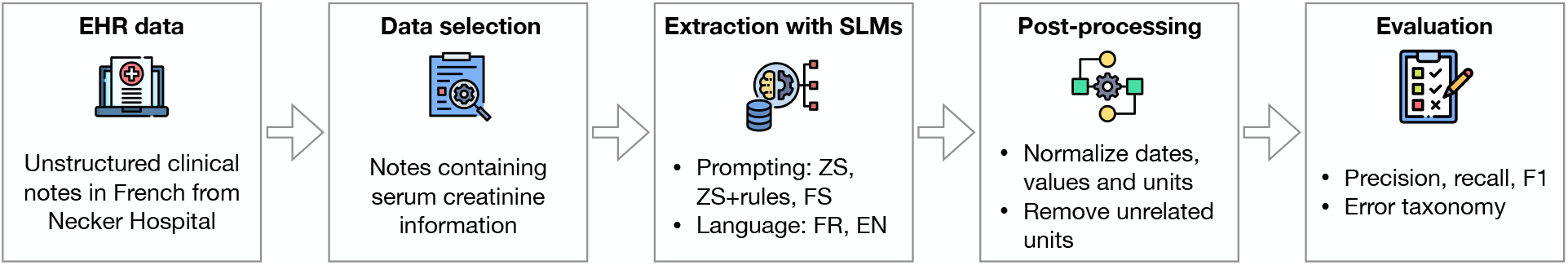
Overall workflow of extracting longitudinal serum creatinine from French clinical notes SLM: small language models, ZS: zero-shot, FS: few-shot

### Target biomarker and data sources

The study included patients diagnosed with ciliopathies followed at Necker Hospital, a national reference center for rare diseases in France. We focused on extracting longitudinal serum creatinine data from clinical notes. Each measurement was defined as a triplet of test date, numerical value, and unit.

Clinical data were retrieved from Necker Hospital’s data warehouse, which integrates heterogeneous sources of EHR, including structured laboratory data and unstructured clinical notes in French (22). We focused on narrative notes. To reduce noise and computational burden, we preselected documents with co-occurrence of the term *creatinine*, a numeric value and a measurement unit (restricted to the two conventional formats used in clinical practice: µmol/L and mg/dL). Notes after kidney failure or renal replacement therapy were excluded as our objective was to reconstruct pre-failure trajectories.

### Small language model selection

We selected open-source SLMs with up to 10 billion parameters, a commonly used threshold for small models, to enable efficient and local deployment. Candidate models were required to generate structured output and to process French-language input. To facilitate comparison across architectures and parameter sizes, we included four recent open-weight models: Mistral-7B-Instruct-v0.3 (15), Llama-3.2-3B-Instruct (16), Qwen3-4B-Instruct, and Qwen3-8B-Instruct (17).

### Preliminary experiments and observed challenges

Preliminary zero-shot experiments revealed multiple challenges in extracting serum creatinine triplets (date, value, unit) from clinical notes, involving both the individual components and incorrect extractions of unrelated tests.

- Date extraction. Dates appeared in heterogeneous formats, including French numeric formats (day/month/year), international formats (year-month-day), and mixed forms with month names. Some retrospective descriptions reported only the year and month, which models extracted as partial dates. Relative expressions such as “ce jour” (“this day” in French) were inconsistently handled: sometimes preserved verbatim, sometimes substituted with the visit date mentioned in the note.
- Value and unit extraction. Numeric values were occasionally returned together with the unit (e.g., “145 µmol/L”), and decimal commas (standard in French) were sometimes preserved as commas. Units were expressed with inconsistent notations (e.g., µ, u, micro), requiring normalization.
- Incorrect extractions. Models extracted other laboratory tests, such as eGFR or urinary creatinine, leading to false positives. The corresponding unit was correctly extracted but did not match serum creatinine. These inconsistencies made these outputs relatively easy to identify and discard. In addition, since most patients were children, clinical notes occasionally mentioned test results from their parents; these values were sometimes erroneously extracted by the models.

These observations motivated the design of targeted prompting strategies and post-processing steps to improve triplet extraction accuracy.

## Experimental design

### SLM prompting

To systematically evaluate the performance of SLMs, we varied two main dimensions: prompting strategy and prompt language. Building on the observed challenges, we tested three prompting strategies: (i) zero-shot, (ii) zero-shot with explicit rules (restricting extraction to serum creatinine, excluding family members’ results, and handling partial or relative dates), and (iii) few-shot (2-shot) with rules, including two representative annotated examples. Prompts were provided in French (the original language of the notes) and English to assess multilingual capabilities.

All models were executed locally with a temperature fixed at 0, using schema-constrained generation within the DSPy framework, which supports declarative prompt programming and structured output generation (23).

For comparison, a regular expression-based extractor was implemented as a baseline. It required the mention of creatinine followed by a numerical value and a valid unit. The extracted values were linked to the nearest date according to a priority window, with allowance for heterogeneous date formats.

Prompt templates and details of the pattern-based extractor are provided in the Supplement.

### Post-processing

To further address the inconsistencies observed in preliminary experiments, we implemented a post-processing pipeline to standardize outputs and reduce noise. Date normalization ensured consistent formatting, with partial dates (month-only or year-only) completed to the first day of the corresponding period. Numeric values were harmonized to a uniform decimal convention. Units were mapped to a controlled vocabulary, and extractions inconsistent with serum creatinine (e.g., units corresponding to eGFR or urine assays) were discarded.

An illustrative example of raw model output and the corresponding cleaned structured triplets is shown in Figure 2.

**Figure 2.**
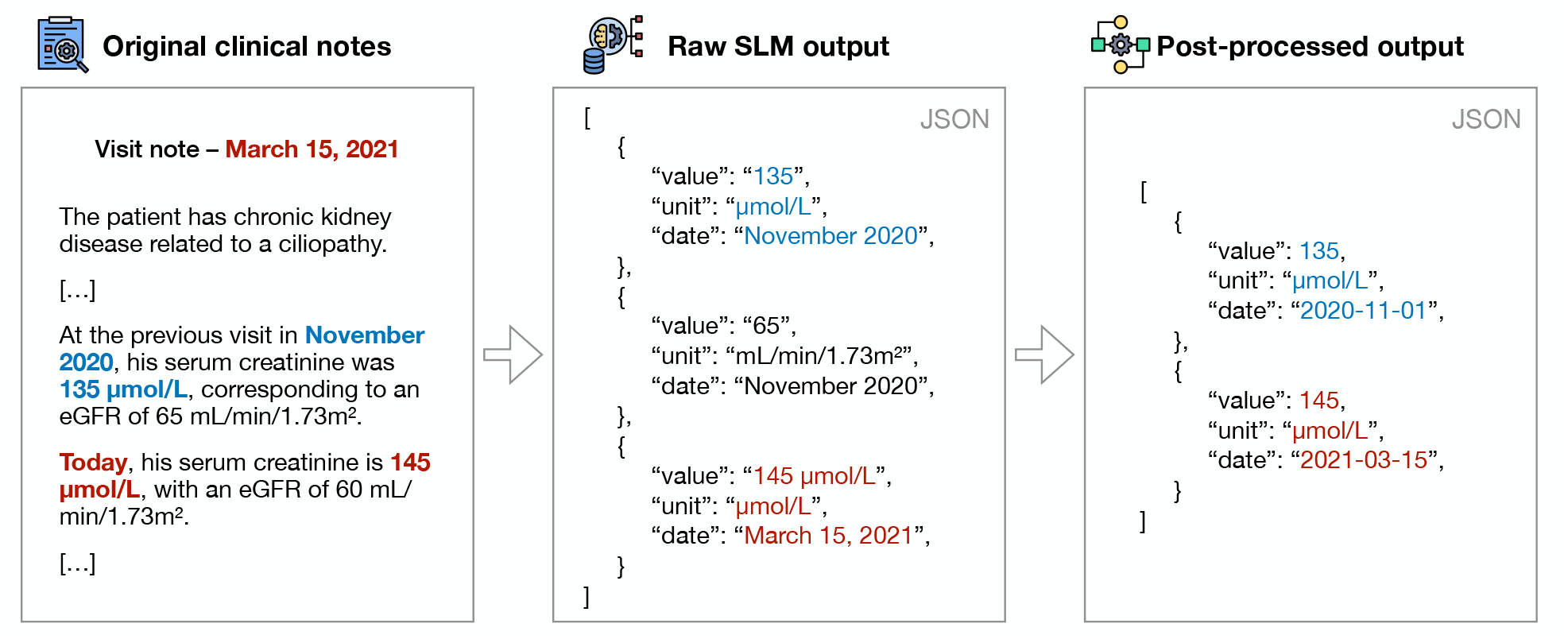
An illustrative example of raw model output and cleaned structured triplets

### Evaluation

Two annotators (XM and CF) independently annotated the triplets (date, value, unit) in each document. Discrepancies were resolved by consensus to establish the final reference standard. Model outputs were then compared against this ground truth. Performance was assessed at the document level, with an extracted triplet considered as correct if all three components matched the ground truth. Precision (the proportion of extracted triplets that were correct), recall (the proportion of ground truth triplets that were successfully retrieved) and F1-score were reported as primary evaluation metrics.

Evaluation of dates allowed a 30-day tolerance window, acknowledging that relative or approximate expressions (e.g., “today”) may not always align exactly with the reference date but remain clinically acceptable for reconstructing longitudinal trajectories over years.

The rule-based baseline was evaluated under the same conditions to contextualize model performance.

## Results

### Dataset characteristics

We analyzed a total of 81 clinical notes from 11 patients with ciliopathies, corresponding to 200 serum creatinine triplets annotated as the ground truth.

As shown in Table 1, token counts varied widely and creatinine measurements were highly skewed, reflecting heterogeneous documentation from brief visit summaries to extensive retrospective narratives. Each note contained more date mentions (mean 20.4) than creatinine measurements (mean 2.5); most dates were absolute, while relative expressions such as *“today”* were less common. This discrepancy highlights the challenge for SLMs to accurately link each measurement to its corresponding date.

**Table 1:**
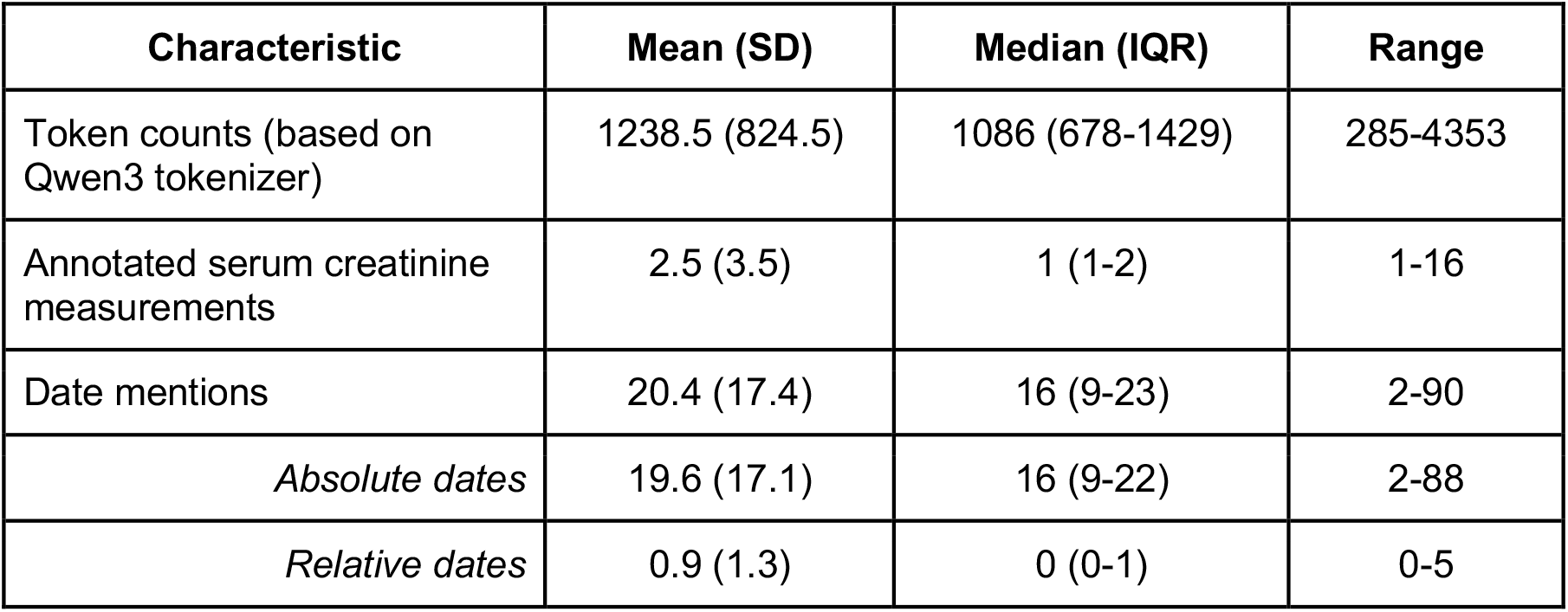
Characteristics of the clinical notes.

| Characteristic | Mean (SD) | Median (IQR) | Range |
| --- | --- | --- | --- |
| Token counts (based on Qwen3 tokenizer) | 1238.5 (824.5) | 1086 (678-1429) | 285-4353 |
| Annotated serum creatinine measurements | 2.5 (3.5) | 1 (1-2) | 1-16 |
| Date mentions | 20.4 (17.4) | 16 (9-23) | 2-90 |
| *Absolute dates* | 19.6 (17.1) | 16 (9-22) | 2-88 |
| *Relative dates* | 0.9 (1.3) | 0 (0-1) | 0-5 |

### Overall extraction performance

Extraction performance across all settings is summarized in Figure 3.

**Figure 3.**
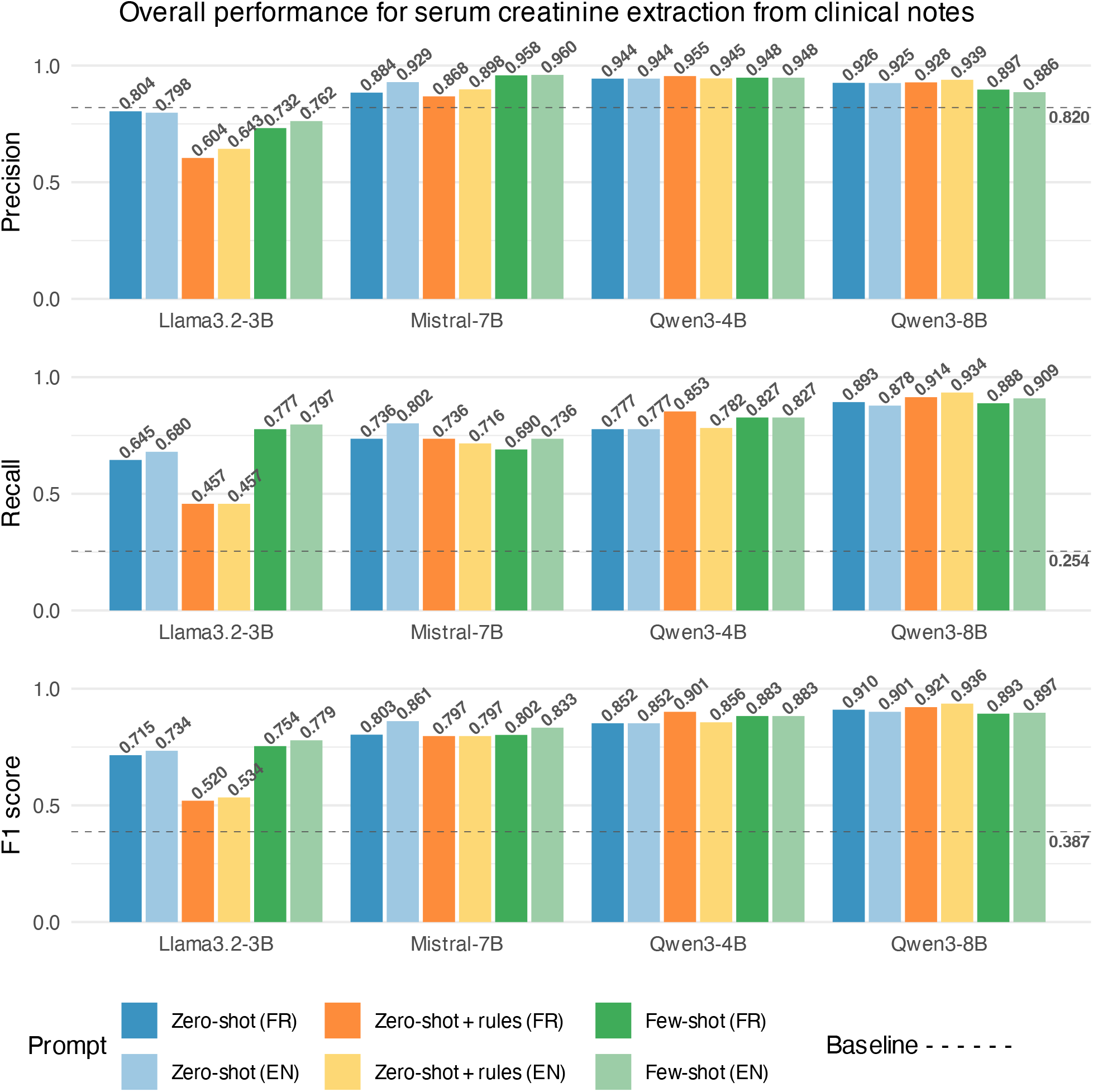
Overall performance

The regex baseline achieved a precision of 0.820, but recall was only 0.254, resulting in an F1-score of 0.387. This confirms that while rule-based extraction can accurately identify values matching predefined patterns, real-world clinical narratives are more complex, with ambiguous dates, missing units, or nonstandard reporting formats that such rules cannot capture. All SLMs successfully extracted structured serum creatinine triplets, with F1-scores ranging from 0.520 (Llama-3.2-3B, zero-shot + rules, FR) to 0.936 (Qwen3-8B, zero-shot + rules, EN). Precision exceeded that of the regex baseline for all models except Llama-3.2-3B, while recall was consistently higher across all SLMs.

Performance varied across models of different sizes. Within the Qwen3 family, the 8B model consistently outperformed the 4B model. Both Qwen3 models outperformed Mistral-7B under most prompting conditions, while Llama-3.2-3B showed the lowest performance overall (F1≤0.779). These findings suggest that increasing model size can improve extraction performance.

We next examined prompting effects. For Mistral-7B, zero-shot English was the best (F1=0.861), adding rules reduced performance, while few-shot increased precision (to 0.960) at the cost of recall. Llama-3.2-3B was penalized by rules (F1 0.520-0.534), while few-shot gave the best within-model performance (F1 0.754-0.779). Qwen3-4B benefited from zero-shot with rules in French (F1=0.901), whereas Qwen3-8B peaked with the same strategy in English (F1=0.936). Overall, rules tended to help Qwen3 models, few-shot was useful for Llama, and Mistral favored simple zero-shot.

Finally, we evaluated the effect of prompt language. Differences between French and English prompts were modest and model-specific. Mistral-7B and Llama-3.2-3B tended to perform better with English prompts, whereas Qwen3 models showed mixed results: Qwen3-4B favored French with rules, while Qwen3-8B peaked with English (F1=0.936). Overall, English had a slight advantage, but French prompts also achieved competitive performance, indicating effective multilingual handling of the tested models.

### Error analysis

We reviewed the post-processed outputs from the best-performing configuration (Qwen-8B, zero-shot with rules in English) to identify the main sources of false positives. Among the 15 false positives, three were incorrect attributions to family members despite the exclusion rule, and twelve were date misclassifications, typically occurring when multiple dates appeared in the same paragraph and the model failed to select the correct one.

We also reviewed the raw outputs to better understand the impact of different prompts. Despite explicit prompt rules, some unrelated tests, most often eGFR, were still extracted in the raw outputs. These were systematically removed during post-processing and therefore did not affect the final performance.

Finally, we examined the models’ ability to consistently process duplicate text. We selected a sentence describing the disease history that included six creatinine measurements and was repeated verbatim across nine notes. Only the best-performing configuration (Qwen-8B, zero-shot with rules in English) correctly extracted all instances, whereas all other models did not achieve full consistency; for example, Mistral-7B with the same prompt succeeded in only five of the nine notes. This indicates that the models differed in their robustness when processing duplicated content in real-world clinical narratives.

## Discussion

In this study, we demonstrated that locally deployed SLMs can effectively extract longitudinal serum creatinine measurements from French clinical notes in the context of rare kidney diseases. By framing the task as the extraction of structured triplets (date, value, unit) and applying targeted post-processing, we achieved high performance across multiple open-source models. The best configuration, Qwen-8B with few-shot prompting and explicit rules, reached an F1-score of 0.928, confirming that SLMs can capture information that pattern-based methods systematically miss in complex real-world notes. Performance varied across models and prompting strategies. Model performance increased with size, with Qwen-8B performing best overall. French and English prompts achieved comparable performance across different SLMs, while prompt strategies were model-specific. Finally, when assessing stability on duplicated content, only Qwen-8B achieved perfect consistency across repeated notes, whereas other models showed variable behavior.

### Comparison with prior work

The use of LLMs for clinical information extraction has expanded recently, and attention has shifted towards small or lightweight models that can be deployed locally while maintaining competitive performance. Promising results have been obtained across diverse domains, including the extraction of echocardiographic entities, classification and information extraction for acute heart failure from French clinical notes, renal histopathology annotation in lupus nephritis, and temporal relation extraction in oncology (24–28). In the rare diseases, hybrid pipelines combining dictionary-based tools with SLMs have also been explored for patient identification (7). To our knowledge, few works have evaluated SLMs for extracting laboratory results as structured triplets across repeated encounters, and none have applied them to the longitudinal reconstruction of biomarkers.

### Clinical and technical significance and strengths

Real-world data are increasingly recognized as an essential complement to rare disease registries, which often suffer from small cohorts and incomplete follow-up (2). In nephrology, studies reconstructing CKD trajectories from EHRs have highlighted major challenges such as irregular testing frequency, missing values, and attrition bias (29). Multiple measurements per year, typically two in clinical follow-up, are needed to capture this variability and model longterm trajectories (30). By enriching structured databases with additional measurements extracted from free-text notes, our pipeline increases the density and continuity of biomarker trajectories and thus enables more complete and reliable longitudinal analyses.

Beyond clinical utility, SLMs offer practical advantages for hospital environments: they are privacy-preserving, energy-efficient, and computationally affordable (10). We demonstrated these benefits in a real-world use case, using French clinical notes from ciliopathy patients. Our results show that lightweight open-source models, when coupled with task-specific prompting and post-processing, can achieve high precision and recall without external data or fine-tuning.

### Documentation quality and data usability

The effectiveness of automated extraction also depends on clinical documentation quality. Growing workload and limited clerical support have led clinicians to produce shorter notes with frequent abbreviations and missing units of lab test results, which hinder automated processing. Conversely, the rise of speech recognition and AI-assisted note generation may re-introduce more detailed narratives. Such verbose but information-rich documentation could greatly enhance the ability of language models to extract clinically meaningful longitudinal data. Maintaining minimal documentation standards that balance efficiency and clarity, such as consistent measurement units, standard abbreviations, and temporal references, can significantly improve downstream data usability. Besides, collaboration between clinicians and informatics specialists will be essential to ensure that new documentation workflows remain compatible with automated data extraction and to develop more intelligent EHR systems with intrinsic coherence checks.

### Limitations

This study has several limitations. First, the dataset was relatively small, with documents from only eleven patients, reflecting the rarity of ciliopathies and the difficulty of building large annotated corpora in rare disease research. Second, the data originated from a single center, which may limit generalizability since documentation style can vary across hospitals. Third, the preselection of notes mentioning “creatinine” with explicit values and units may have excluded incomplete or misspelled entries, thereby lowering task difficulty. Fourth, the design of our SLM prompts was guided by preliminary experiments on this dataset, potentially introducing adaptations specific to the reporting patterns in these documents and limiting transferability. Finally, although SLMs occasionally extracted related tests such as eGFR, we excluded these outputs during post-processing and thus did not fully assess their ability to jointly extract and differentiate closely related biomarkers.

### Future directions

Renal function is a prototypical example where longitudinal biomarker trajectories are essential for monitoring disease progression. Future work should further evaluate the ability of SLMs to support renal monitoring by extracting a broader set of kidney-related variables (e.g. proteinuria, electrolytes) and by building more comprehensive longitudinal datasets. This is particularly important in rare diseases, where the limited number of patients must be compensated by a “more data per case” paradigm. Beyond renal disease, the same approach could generalize to other chronic conditions, such as lipid profiles for cardiovascular risk, liver enzymes for chronic liver disease, and creatine kinase for neuromuscular disorders. We also aim to capture temporal attributes for phenotype entities, thereby generating longitudinal patient histories from EHR narratives. Another direction is to improve subject attribution (e.g., distinguishing patients from family members), which may require advanced prompting or task-specific fine-tuning. Finally, systematic comparisons between SLMs and LLMs are needed to clarify trade-offs between performance, resource demands, and scalability in real-world clinical settings.

## Conclusion

In this study, we developed and validated an SLM-based pipeline to extract longitudinal serum creatinine measurements from French clinical notes of patients with renal ciliopathies. The models achieved consistently high performance, with Qwen-8B reaching an F1-score of 0.928. By coupling lightweight language models with targeted prompting and post-processing, our approach effectively transforms unstructured narratives into analyzable data. This method not only enhances the completeness of real-world EHR datasets but also addresses a major challenge in rare kidney disease research, where longitudinal information is crucial yet often missing. Extending this framework to additional biomarkers and diseases will further establish SLMs as practical, privacy-preserving tools for clinical research and precision medicine.

## Supporting information

Prompt templates; Regular expression baseline extractor

## Data Availability

All data produced in the present study are available upon reasonable request to the authors

## Ethical Approval Statement

The study was conducted within the framework of the TheRaCil consortium (EU Horizon Programme, grant agreement No. 101080717) and was approved by the French Ethics and Scientific Committee for Health Research (CESREES, reference 2201437).

## References

1. The Lancet Global Health. The landscape for rare diseases in 2024. Lancet Glob Health. 2024 Mar;12(3):e341.

2. Adang LA, Sevagamoorthy A, Sherbini O, Fraser JL, Bonkowsky JL, Gavazzi F, et al. Longitudinal natural history studies based on real-world data in rare diseases: Opportunity and a novel approach. Mol Genet Metab. 2024 May 1;142(1):108453.

3. Wang Y, Wang L, Rastegar-Mojarad M, Moon S, Shen F, Afzal N, et al. Clinical Information Extraction Applications: A Literature Review. J Biomed Inform. 2018 Jan;77:34–49.

4. Bedi S, Liu Y, Orr-Ewing L, Dash D, Koyejo S, Callahan A, et al. Testing and Evaluation of Health Care Applications of Large Language Models: A Systematic Review. JAMA. 2025 Jan 28;333(4):319–28.

5. Gu B, Shao V, Liao Z, Carducci V, Brufau SR, Yang J, et al. Scalable information extraction from free text electronic health records using large language models. BMC Med Res Methodol. 2025 Jan 28;25(1):23.

6. Richter-Pechanski P, Seiferling M, Kiriakou C, Schwab DM, Geis NA, Dieterich C, et al. Medication information extraction using local large language models. J Biomed Inform. 2025 Sept;169:104898.

7. Wu J, Dong H, Li Z, Wang H, Li R, Patra A, et al. A hybrid framework with large language models for rare disease phenotyping. BMC Med Inform Decis Mak. 2024 Oct 8;24(1):289.

8. Ntinopoulos V, Rodriguez Cetina Biefer H, Tudorache I, Papadopoulos N, Odavic D, Risteski P, et al. Large language models for data extraction from unstructured and semi-structured electronic health records: a multiple model performance evaluation. BMJ Health Care Inform. 2025 Jan 19;32(1):e101139.

9. Hallowell N, Parker M, Nellåker C. Big data phenotyping in rare diseases: some ethical issues. Genet Med. 2019 Feb;21(2):272–4.

10. Garg M, Raza S, Rayana S, Liu X, Sohn S. The Rise of Small Language Models in Healthcare: A Comprehensive Survey [Internet]. arXiv; 2025 [cited 2025 July 8]. Available from: http://arxiv.org/abs/2504.17119

11. Wang F, Zhang Z, Zhang X, Wu Z, Mo T, Lu Q, et al. A Comprehensive Survey of Small Language Models in the Era of Large Language Models: Techniques, Enhancements, Applications, Collaboration with LLMs, and Trustworthiness [Internet]. arXiv; 2024 [cited 2025 Oct 9]. Available from: http://arxiv.org/abs/2411.03350

12. Corradini F, Leonesi M, Piangerelli M. State of the Art and Future Directions of Small Language Models: A Systematic Review. Big Data Cogn Comput. 2025 July;9(7):189.

13. Kim H, Hwang H, Lee J, Park S, Kim D, Lee T, et al. Small language models learn enhanced reasoning skills from medical textbooks. Npj Digit Med. 2025 May 2;8(1):240.

14. Belcak P, Heinrich G, Diao S, Fu Y, Dong X, Muralidharan S, et al. Small Language Models are the Future of Agentic AI [Internet]. arXiv; 2025 [cited 2025 Oct 13]. Available from: http://arxiv.org/abs/2506.02153

15. Jiang AQ, Sablayrolles A, Mensch A, Bamford C, Chaplot DS, Casas D de las, et al. Mistral 7B [Internet]. arXiv; 2023 [cited 2025 Sept 17]. Available from: http://arxiv.org/abs/2310.06825

16. Grattafiori A, Dubey A, Jauhri A, Pandey A, Kadian A, Al-Dahle A, et al. The Llama 3 Herd of Models [Internet]. arXiv; 2024 [cited 2025 Sept 17]. Available from: http://arxiv.org/abs/2407.21783

17. Yang A, Li A, Yang B, Zhang B, Hui B, Zheng B, et al. Qwen3 Technical Report [Internet]. arXiv; 2025 [cited 2025 Sept 17]. Available from: http://arxiv.org/abs/2505.09388

18. Aymé S, Bockenhauer D, Day S, Devuyst O, Guay-Woodford LM, Ingelfinger JR, et al. Common Elements in Rare Kidney Diseases: Conclusions from a Kidney Disease: Improving Global Outcomes (KDIGO) Controversies Conference. Kidney Int. 2017 Oct 1;92(4):796–808.

19. Carrero JJ, Fu EL, Vestergaard SV, Jensen SK, Gasparini A, Mahalingasivam V, et al. Defining measures of kidney function in observational studies using routine health care data: methodological and reporting considerations. Kidney Int. 2023 Jan;103(1):53–69.

20. Petzold F, Billot K, Chen X, Henry C, Filhol E, Martin Y, et al. The genetic landscape and clinical spectrum of nephronophthisis and related ciliopathies. Kidney Int. 2023 Aug;104(2):378–87.

21. Wang X, Faviez C, Douillet M, Knebelmann B, Garcelon N, Burgun A, et al. Enhancing Longitudinal Data Analysis with Unstructured EHRs: A Case Study of Renal Function Evaluation in Rare Disease. Stud Health Technol Inform. 2025 May 15;327:1270–4.

22. Garcelon N, Neuraz A, Salomon R, Faour H, Benoit V, Delapalme A, et al. A clinician friendly data warehouse oriented toward narrative reports: Dr. Warehouse. J Biomed Inform. 2018 Apr 1;80:52–63.

23. Khattab O, Singhvi A, Maheshwari P, Zhang Z, Santhanam K, Vardhamanan S, et al. DSPy: Compiling Declarative Language Model Calls into Self-Improving Pipelines [Internet]. arXiv; 2023 [cited 2025 Sept 16]. Available from: http://arxiv.org/abs/2310.03714

24. Huang J, Yang DM, Rong R, Nezafati K, Treager C, Chi Z, et al. A critical assessment of using ChatGPT for extracting structured data from clinical notes. Npj Digit Med. 2024 May 1;7(1):106.

25. Bazoge A, Wargny M, Constant dit Beaufils P, Morin E, Daille B, Gourraud PA, et al. Assessing large language models for acute heart failure classification and information extraction from French clinical notes. Comput Biol Med. 2025 Sept 1;195:110609.

26. Chi J, Rouphail Y, Hillis E, Ma N, Nguyen A, Wang J, et al. EchoLLM: extracting echocardiogram entities with light-weight, open-source large language models. JAMIA Open. 2025 Aug;8(4):ooaf092.

27. Tai ICY, Wong ECK, Wu JT, Leung K, Yap DYH, Wong ZSY. Exploring Offline Large Language Models for Clinical Information Extraction: A Study of Renal Histopathological Reports of Lupus Nephritis Patients. Stud Health Technol Inform. 2024 Aug 22;316:899–903.

28. Sharma V, Fernandez A, Ioanovici A, Talby D, Buijs F. Lexicans at Chemotimelines 2024: Chemotimeline Chronicles - Leveraging Large Language Models (LLMs) for Temporal Relations Extraction in Oncological Electronic Health Records. In: Naumann T, Ben Abacha A, Bethard S, Roberts K, Bitterman D, editors. Proceedings of the 6th Clinical Natural Language Processing Workshop [Internet]. Mexico City, Mexico: Association for Computational Linguistics; 2024 [cited 2025 Oct 2]. p. 394–405. Available from: https://aclanthology.org/2024.clinicalnlp-1.38/

29. Cleary F, Prieto-Merino D, Hull S, Caplin B, Nitsch D. Feasibility of evaluation of the natural history of kidney disease in the general population using electronic healthcare records. Clin Kidney J. 2021 June 1;14(6):1603–9.

30. Wong K, Pitcher D, Braddon F, Downward L, Steenkamp R, Annear N, et al. Effects of rare kidney diseases on kidney failure: a longitudinal analysis of the UK National Registry of Rare Kidney Diseases (RaDaR) cohort. The Lancet. 2024 Mar;403(10433):1279–89.

