## Supplementary material for "Longitudinal information extraction from clinical notes in rare diseases: an efficient approach with small language models": Prompt templates; Regular expression baseline extractor

#### S1. Prompt templates

The following prompt templates were used in our experiments to evaluate different prompting strategies (zero-shot, zero-shot with rules, and few-shot with rules). Each template was tested in both French and English.

##### Prompt 1 – Zero-shot (French)

Extraire les mesures structurées de la créatinine sérique à partir d’un texte clinique en français.

##### Prompt 1 – Zero-shot (English)

Extract structured serum creatinine measurements from the following French clinical text.

##### Prompt 2 – Zero-shot with rules (French)

Extraire toutes les mesures de créatinine sérique dans le texte suivant.

Pour chaque mesure, donner un objet JSON avec :
- value : la valeur numérique
- unit : "µmol/L" ou "mg/dL"
- date : la date associée, si elle existe

Règles :
1. Ne garder que les mesures de créatinine dans le sang. Ne pas extraire la créatinine urinaire, la clairance ou le DFG (eGFR).
2. Si la date est "ce jour" ou "aujourd’hui", utiliser la date de consultation si elle est présente. Sinon, garder "ce jour".
3. Si la date ne contient qu’un mois ou une année (ex : "mars 2020", "2020"), la garder telle quelle.
4. Ne pas extraire les résultats de créatinine d’un membre de la famille (père, mère, frère, etc.).

##### Prompt 2 – Zero-shot with rules (English)

Extract all serum creatinine measurements from the following text.

For each measurement, return a JSON object with:
- value: the numeric value
- unit: either "µmol/L" or "mg/dL"
- date: the associated date, if available

Rules:
1. Only extract creatinine in blood. Do not include urine creatinine, clearance, or eGFR.
2. If the date is "ce jour" or "aujourd'hui", use the consultation date if present. Otherwise, keep "ce jour".
3. If the date includes only a month or year (e.g., "mars 2020", "2020"), keep it as written.
4. Do not extract creatinine results that belong to a family member (e.g., father, mother, sibling).

#### Prompt 3 – Few-shot with rules (French)

Extraire toutes les mesures de créatinine sérique dans le texte suivant.

Pour chaque mesure, donner un objet JSON avec :
- value : la valeur numérique
- unit : "µmol/L" ou "mg/dL"
- date : la date associée, si elle existe

Règles :
1. Ne garder que les mesures de créatinine dans le sang. Ne pas extraire la créatinine urinaire, la clairance ou le DFG (eGFR).
2. Si la date est "ce jour" ou "aujourd'hui", utiliser la date de consultation si elle est présente. Sinon, garder "ce jour".
3. Si la date ne contient qu'un mois ou une année (ex : "mars 2020", "2020"), la garder telle quelle.
4. Ne pas extraire les résultats de créatinine d'un membre de la famille (père, mère, frère, etc.).

Voici deux exemples :

Texte :
"xxx"
Sortie :
[{"value": "xxx", "unit": "µmol/L", "date": "xxx"}]

Texte :
"xxx"
Sortie :
[
  {"value": "xxx", "unit": "µmol/L", "date": "xxx"},

{"value": "xxx", "unit": "µmol/L", "date": "xxx"}
]

##### Prompt 3 – Few-shot with rules (English)

Extract all serum creatinine measurements from the following text.

For each measurement, return a JSON object with:
- value: the numeric value
- unit: either "µmol/L" or "mg/dL"
- date: the associated date, if available

Rules:
1. Only extract creatinine in blood. Do not include urine creatinine, clearance, or eGFR.
2. If the date is "ce jour" or "aujourd'hui", use the consultation date if present. Otherwise, keep "ce jour".
3. If the date includes only a month or year (e.g., "mars 2020", "2020"), keep it as written.
4. Do not extract creatinine results belonging to a family member (e.g., father, mother, sibling).

Here are two examples:

Text:
"xxx"
Output:
[{"value": "xxx", "unit": "µmol/L", "date": "xxx"}]

Text:
"xxx"
Output:
[
{"value": "xxx", "unit": "µmol/L", "date": "xxx"},
{"value": "xxx", "unit": "µmol/L", "date": "xxx"}
]

### S2. Regular expression baseline extractor

###### Anchor pattern

- The regex searches for creatinine keywords (e.g., créatinine, créatininémie, créat) followed by a numerical value and a required unit.
- Short connectors or descriptive phrases (up to 30 characters, e.g., *est à*, *revient à*) are tolerated between the keyword and the value.

- Accepted units are strictly limited to µmol/L, umol/L, micromol/L, and mg/dL.

**Confounder exclusion**

- Mentions are discarded if, within a ±40 character window, the text refers to another test containing the word "creatinine", such as urinary creatinine, albumin/creatinine ratio (ACR), or creatinine clearance.

**Date linking**

- Each extracted value is linked to the closest date token according to the following priority:
    1. Medium-distance window before the mention (≤800 characters).
    2. Near-distance window after the mention (≤200 characters).
    3. Far-distance window before the mention (≤3000 characters).
    4. If no date is found, the field is left missing (NA).
- Recognized date formats include ISO (YYYY-MM-DD), numeric French (dd/mm/yyyy), textual French (e.g., *12 avril 2021*), month–year, year-only (*en 2001*), and relative terms (*ce jour*, *aujourd'hui*).
